# Detection of malignant peripheral nerve sheath tumors in patients with neurofibromatosis using aneuploidy and mutation identification in plasma

**DOI:** 10.1101/2021.09.09.21262860

**Authors:** Austin K. Mattox, Christopher Douville, Natalie Silliman, Janine Ptak, Lisa Dobbyn, Joy Schaefer, Maria Popoli, Cherie Blair, Kathy Judge, Kai Pollard, Christine Pratilas, Jaishri Blakeley, Fausto Rodriguez, Allan Belzberg, Chetan Bettegowda

## Abstract

Malignant peripheral nerve sheath tumors (MPNST) are the deadliest cancer that arises in individuals diagnosed with neurofibromatosis and account for nearly 5% of the 15,000 soft tissue sarcomas diagnosed in the United States each year. Comprised of neoplastic Schwann cells, primary risk factors for developing MPNST include existing plexiform neurofibromas (PN), prior radiotherapy treatment, and expansive germline mutations involving the entire *NF1* gene and surrounding genes. PN develop in nearly 30-50% of patients with NF1 and most often grow rapidly in the first decade of life. One of the most important aspects of clinical care for NF1 patients is monitoring PN for signs of malignant transformation to MPNST that occurs in 10-15% of patients.

We perform aneuploidy analysis on ctDNA from 883 ostensibly healthy individuals and 28 patients with neurofibromas, including 7 patients with benign neurofibroma, 9 patients with PN and 12 patients with MPNST. Overall sensitivity for detecting MPNST using genome wide aneuploidy scoring was 33%, and analysis of sub-chromosomal copy number alterations (CNAs) improved sensitivity to 50% while retaining a high specificity of 97%. In addition, we performed mutation analysis on plasma cfDNA for a subset of patients and identified mutations in *NF1, NF2, RB1, TP53BP2*, and *GOLGA2*. Given the high throughput and relatively low sequencing coverage required by our assay, liquid biopsy represents a promising technology to identify incipient MPNST.

## INTRODUCTION

Neurofibromatosis type 1 (NF1) is caused by inherited or *de novo* mutations in the *NF1* gene that codes for the cytoplasmic protein neurofibromin^1^. Neurofibromin is a GTPase-activating protein (GAP) for the RAS family of proto-oncogenes, and mutations in *NF1* lead to persistent RAS signaling and uncontrolled cellular growth through downstream RAF, MEK, and ERK signalling^1,2^. Activated RAS resulting from the loss of GTPase activity of NF1 also leads to downstream activation of the PI3K/AKT/mTOR pathway, further contributing to increased proliferation^2^.

Malignant peripheral nerve sheath tumors (MPNST) are the deadliest cancer that arises in individuals diagnosed with NF1 and account for nearly 5% of the 15,000 soft tissue sarcomas diagnosed in the United States each year^3^. Comprised of neoplastic Schwann cells, primary risk factors for developing MPNST include existing plexiform neurofibromas (PN), prior radiotherapy treatment, and expansive germline mutations involving the entire *NF1* gene and surrounding genes^4^. PN develop in nearly 30-50% of patients with NF1 and most often grow rapidly in the first decade of life. One of the most important aspects of clinical care for NF1 patients is monitoring PN for signs of malignant transformation to MPNST that occurs in 10-15% of patients.

Bi-allelic loss of *NF1* is not sufficient for malignant transformation of PN to MPNST^5-7^. Additional mutations or copy number alterations of genes such as *TP53, SUZ12, EGFR, CDKN2A*, and *TERT* that are often not present in benign PN suggest that these alterations represent advanced progression to atypical neurofibroma (AN) and MPNST^8-12^.

Data from National Cancer Institute (NCI) NF1 Natural History Study suggest that nearly 50% of patients with PN develop well-demarcated nodular areas within the PN that are larger than 3 cm in size, lack the central dot sign characteristic of PN, and typically show more rapid growth^13^. These distinct nodular lesions (DNL) correlate with pain, and biopsy/resection of DNL leads to a confirmed diagnosis of AN in 70% of cases^14^. Of all confirmed AN cases in the study, all were DNL by MRI and were associated with a modest FDG update of [SUV] = 2.7.

Despite these preliminary results, MRI is unable to reliably differentiate between benign and malignant tumors^15^. Additional studies have suggested FDG-PET has sensitivities of nearly 90% in symptomatic patients, but only when using an SUV cutoff of 3.5 and a non-standard clinical protocol of delayed imaging at 4 hours^16^. Using similar criteria, FDG-PET may have similar sensitivities for monitoring asymptomatic patients for malignant transformation, but only at 49.5% specificity^17^. Pathologically, there are no standardized pathognomonic genetic alterations or immunohistochemical stains to differentiate MPNST from other sarcomas. While gross specimens that clearly arise from nerves lend credence to a diagnosis of MPNST, negative staining for cytokeratins and melanoma markers like Melan-A, MITF, and HMB45 can be useful in distinguishing MPNST from carcinoma and melanoma^18-20^. S100 expression is also decreased or completely lost in MPNST^19^. Genomic loss of *NF1* and *CDKN2A* are thought to be lost early in disease progression but testing for these mutations is also not clinically standardized.

Given the lack of specific imaging and pathologic diagnostic criteria to diagnose MPNST, more accurate and cost-effective biomarkers are needed. Liquid biopsies that assay for mutations or aneuploidy in circulating tumor DNA (ctDNA) represent an attractive, minimally invasive option that could be performed at each longitudinal patient visit. Mutations in polycomb repressive complex 2 (PRC2) subunits such as *SUZ12* and *EED* are found in nearly 70% of MPNST^9,12^. Mutations in β-III-spectrin have also been found in up to 90% of MPNST^21^. Additional Ras pathway activating mutations in genes such as *PIK3CA, KIT, PDGFRA, PTPN11, FGFR1*, and RASSF9, and cell-cycle gene mutations in genes such as *RB1* and *CHEK2* have also been described^22^.

Liquid biopsies also have the advantage over traditional biopsies of capturing tumor heterogeneity. This is important because within a single tumor, there may be areas of PN, AN, low grade MPNST, and high grade MPNST, and traditional single-site biopsy may not capture the most malignant site.

In the present study, we perform aneuploidy analysis on 883 ostensibly healthy individuals and 28 patients with neurofibromas, including 7 patients with benign neurofibroma, 9 patients with PN and 12 patients with MPNST. While overall sensitivity for detecting NF using genome wide aneuploidy measurements was limited, analysis of sub-chromosomal changes may be promising for detecting MPNST. In addition, we performed mutation analysis on plasma cfDNA for a subset of patients and identified mutations in *NF1, NF2, RB1, TP53BP2*, and *GOLGA2*.

## RESULTS

### Patient Characteristics

The primary objective of this pilot study was to differentiate MPNST from PN using genome wide and focal aneuploidy analysis of cfDNA isolated from plasma. To quantify the rate of genome wide CNAs detected in plasma cfDNA of healthy persons, we analyzed 883 samples from a previously published study^23^ using a revised RealSeqS algorithm and a median of 10,223,275 UIDs per sample **(Supplemental Table 1)**. Our patient cohort included 28 patients with NF, including 7 patients with neurofibromas, 9 patients with PN, and 12 patients with MPNST, analyzed in the same manner as healthy controls with a median of 11,240762 UIDs per sample **(Supplemental Table 2)**. All samples had matched leukocyte analysis to exclude germline CNAs. For patients with biopsy-confirmed MPNST, 58% (7/12) had positive PET scans, 17% (2/12) had prior chemotherapy, 17% (2/12) had prior chemotherapy and radiation, 8% (1/12) had prior radiation only, 17% (2/12) had prior surgery only, and 8% (1/12) had prior surgery, chemotherapy, and radiation. Median length of follow up was 523 days.

### Analysis of Genome Wide Aneuploidy

Ninety-six percent (27/28) of patients enrolled onto our study met the criteria for NF1 diagnosis. RealSeqS, which amplifies approximately 750,000 loci across 39 chromosome arms, was used to calculate a genome wide aneuploidy score (GAS) to call plasma samples positive or negative at 97% specificity as determined by the 883 healthy controls. The inclusion of the large number of healthy controls is especially important because it allows for a realistic estimation of specificity and a comparison between healthy persons, patients with neurofibromas, as those with MPNST, as would be done in a real-world setting. The median GAS score in healthy controls was 0.008 ± 0.102, and 0.005 ±0.249 and 0.018 ± 0.652 in benign/plexiform neurofibromas and MPNST, respectively **(Figure 1)**. At 97% specificity in healthy controls, at the time of blood draw, the false positive rate among benign/plexiform neurofibromas was 6.3% (1/16, *p* = 0.42 compared to healthy controls), while the sensitivity for detecting MPNST was 33% (4/12, *p* < 0.001 compared to healthy controls). GAS score did not correlate with tumor volume (R^2^ = 0.09), history of prior adjuvant therapy (*p* = 0.64) or PET positivity (*p* = 0.25), but patients who were alive at the time of last follow up had lower GAS scores (*p* = 0.045).

**Figure 1.**
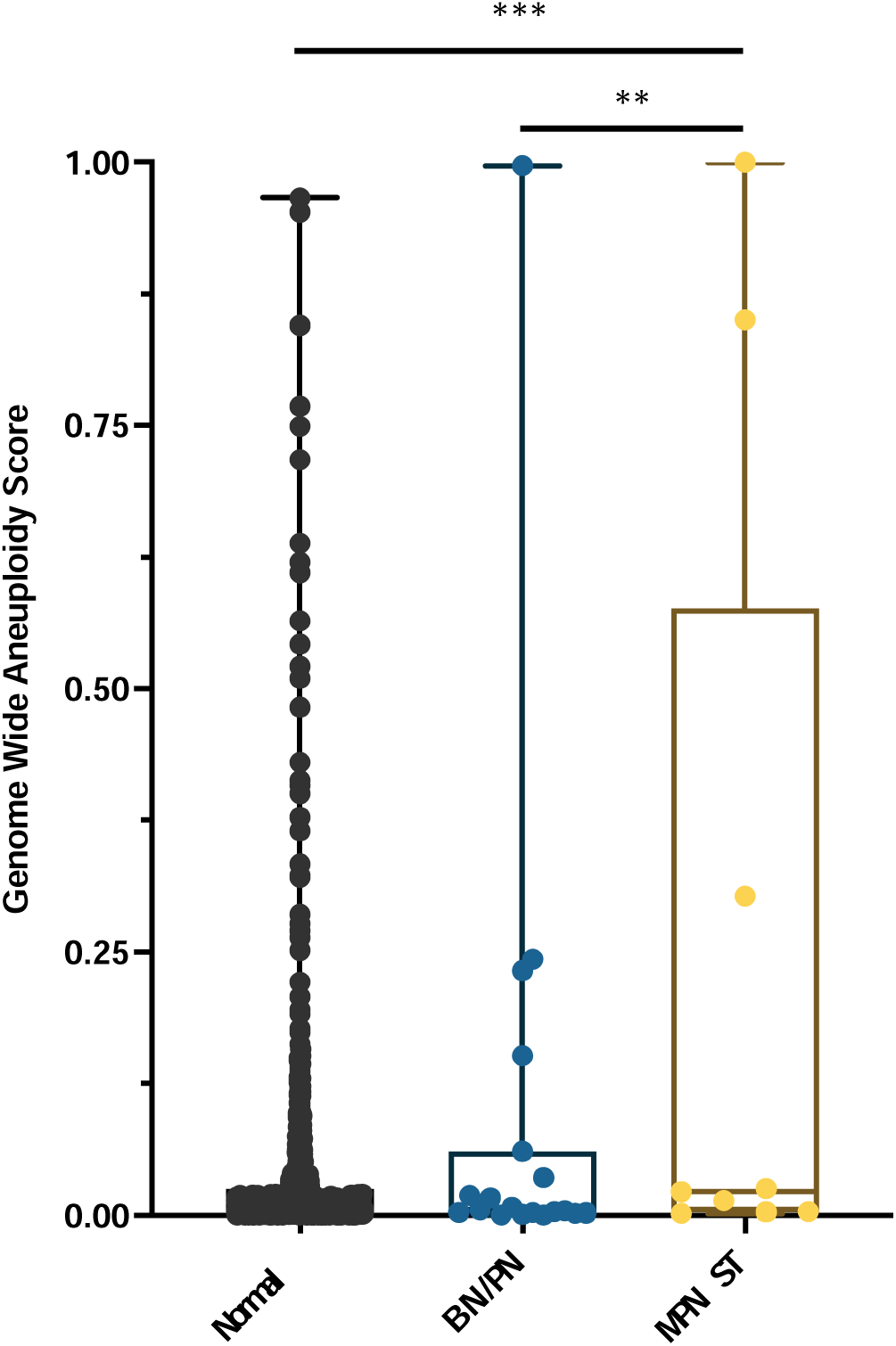
Distribution of genome wide aneuploidy (GAS) scores in healthy individuals and patients with benign (BN) or plexiform neurofibronas (PN) or MPNST. ** *p* < 0.01, *** *p* < 0.001

Interestingly, the one patient with the plexiform neurofibroma deemed to be a false positive at the time of blood draw (INDIA 1283, **Supplemental Table 3**) had a GAS score of 0.997. Biopsy at the time of blood draw confirmed diffuse and atypical changes in the PN. Upon later review, this patient progressed to MPNST 25 months after blood draw, suggesting that aneuploidy analysis significantly pre-dated clinical progression.

### Analysis of Focal Copy Number Alterations

In addition to assessing genome wide aneuploidy, RealSeqS can detect focal amplifications and deletions across 39 chromosome arms. We profiled sub-chromosomal changes across 13 chromosome arms commonly altered in MPNST, including 4q (*PDGFRA*), 5p (*TERT*), 6q (*TBX1*), 7p (*EGFR*), 7q (*BRAF*), 8q (*EXT1*), 9p (*CDKN2A* and *CDKN2B*), 10q (*PTEN*), 11p (*EXT2*), 11q (*EED*), 15q (*IDH2*), 17p (*TP53*), and 17q (*NF1* and *SUZ12*) **(Supplemental Table 4)**. In benign/plexiform NF, only one patient had a focal deletion across all loci assayed. Interestingly, this patient (INDIA 1283) had a deletion in *TERT* and was the same patient that had a GAS score of 0.997 that later progressed to MPNST. Among the 12 patients with MPNST, 17% (2/12) had losses in *TERT*, 8% (1/12) had a loss at *TP53*, and 50% (6/12) had losses on 17q at *SUZ12* **(Figure 2, Supplemental Figure 1)**. These data suggest that focal changes may be useful biomarkers of progression to MPNST.

**Figure 2.**
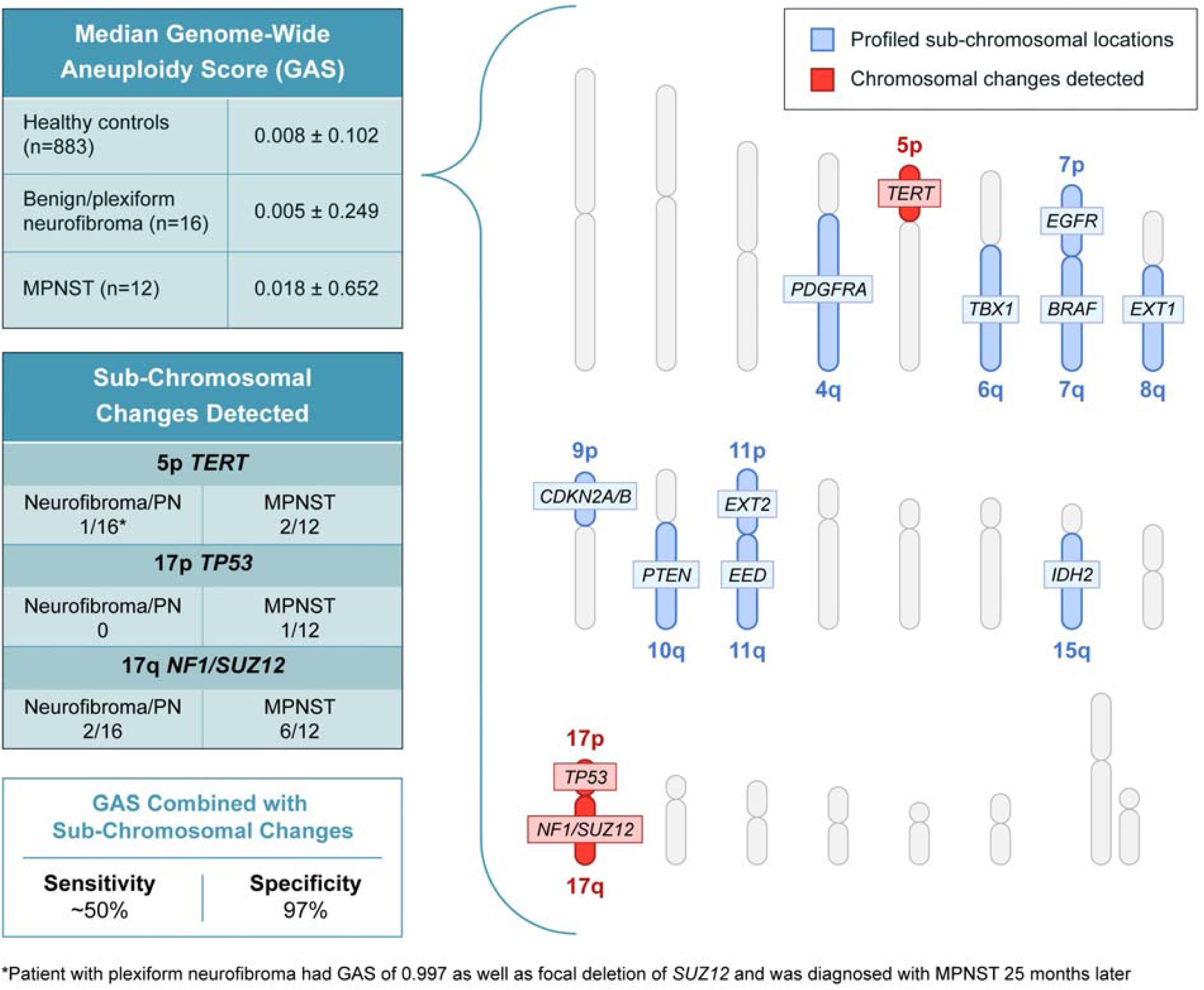
Genome wide aneuploidy scores and detection of sub-chromosomal copy number changes in *PDGFA, TERT, TBX1, EGFR, BRAF, EXT1, CDKN2A/B, PTEN, EXT2, EED, IDH2, TP53, NF1*, and *SUZ12* allow for detection of 50% of MPNST at 97% specificity.

### ctDNA Mutation Analysis

Enough banked plasma was available from 6 patients, 2 with benign neurofibromas and 4 with MPNST, to assay ctDNA for mutations using ddPCR **(Supplemental Tables 1, 5, and 6)**. 3 patients had positive ctDNA results, including one patient (INDIA 1280) with a benign neurofibroma of the right femoral nerve that had a *GOLGA2* splice site acceptor mutation at an MAF of 0.19%, and two MPNST patients (INDIA 1244 and INDIA 1281) that a *RB1* R787Qfs*23 mutation at 2.55% and a *TP53BP2* A324V mutation at 0.04%, respectively.

## DISCUSSION

One of the major clinical challenges in caring for individuals with NF1 is to be able to identify an incipient MPNST. Currently, only anatomic and PET imaging has been shown to be effective^24-26^. Use of PET imaging to track recurrence additionally requires that the primary benign neurofibroma is PET positive before treatment and necessitates the use of radionuclides^24,27^. Thus, alternative methods to monitor disease progression to MPNST, like liquid biopsy, are greatly needed.

To our knowledge, this is the first published study to differentiate MPNST from pre-malignant neurofibromas using aneuploidy and mutation analysis of ctDNA. While our study suggests that genome wide aneuploidy scoring alone may not have high sensitivity for detecting the progression from PN to MPNST, the combination of GAS and detection of sub-chromosomal changes in genes such as *TERT, TP53*, and *SUZ12* or mutations in ctDNA may lead to a sensitivity of ∼50% at a high specificity of 97%. Our sensitivity is likely impacted by low genome wide sequencing coverage, which may be improved by a focused panel that covers the most common copy number alterations (CAN) in MPNST^12,22^.

Malignant transformation of a plexiform neurofibroma to MPSNT can occur over years, and given the mean follow up is 17 months, we cannot definitively identify all cases that may have progressed. In our study, the one patient with PN who had an overwhelmingly positive GAS score of 0.997 and a focal deletion of *SUZ12* at the time of initial blood draw was diagnosed with an MPNST 25 months later.

Main limitations of our study include the relatively small number of patients, the lack of ctDNA data for all patients, and lack of follow up blood draws at fixed intervals. A key strength of our study is the relatively low sequencing coverage needed to detect both genome wide and sub-chromosomal CNAs. The ability to multiplex samples lends high throughput, as each sample only requires ∼10M reads to identify relevant CNAs. This is important for real world implementation as most aneuploidy studies typically utilize 6-10x the amount of sequencing, greatly increasing the cost and limiting feasibility. While our data allow us to make some inferences about CNAs and disease progression from PN to MPNST, it will be important for future prospective studies to collect additional blood samples over a longer period to determine whether GAS, focal CNAs, or ctDNA positivity predicts progression and overall survival.

## MATERIALS AND METHODS

### Patients

All individuals participating in the study provided written informed consent after approval by the institutional review board at The Johns Hopkins IRB00075499. The study complied with the Health Insurance Portability and Accountability Act and the Deceleration of Helsinki.

### Library Construction and Whole Exome Sequencing Buffer and PCR Conditions

Tumor and matched lymphocytic normal DNA library preparation was performed as previously described^28^. Genomic DNA from tumor and normal samples were fragmented and used for Illumina TruSeq library construction (Illumina, San Diego, CA) according to the manufacturer’s instructions. DNA was purified using Agencourt AMPure XP beads (Beckman Coulter, IN) in a ratio of 1.0 to 0.9 of PCR product to beads. Purified, fragmented DNA was mixed with 36 µl of H_2_O, 10 µl of End Repair Reaction Buffer, 5 µl of End Repair Enzyme Mix (cat# E6050, NEB, Ipswich, MA). The 100 µl end-repair mixture was incubated at 20°C for 30 min, and purified using Agencourt AMPure XP beads (Beckman Coulter, IN) in a ratio of 1.0 to 1.25 of PCR product to beads. 42 µl of end-repaired DNA was mixed with 5 µl of 10X dA Tailing Reaction Buffer and 3 µl of Klenow (exo-)(cat# E6053, NEB, Ipswich, MA). The 50 µl mixture was incubated at 37°C for 30 min and purified using Agencourt AMPure XP beads (Beckman Coulter, IN) in a ratio of 1.0 to 1.0 of PCR product to beads. 25 µl of A-tailed DNA was mixed with 6.7 µl of H_2_O, 3.3 µl of PE-adaptor (Illumina), 10 µl of 5X Ligation buffer and 5 µl of Quick T4 DNA ligase (cat# E6056, NEB, Ipswich, MA). The ligation mixture was incubated at 20°C for 15 min and purified using Agencourt AMPure XP beads (Beckman Coulter, IN) in a ratio of 1.0 to 0.95 and 1.0 of PCR product to beads.

To obtain an amplified library, twelve PCRs of 25 µl each were set up, each including 15.5 µl of H_2_O, 5 µl of 5 x Phusion HF buffer, 0.5 µl of a dNTP mix containing 10 mM of each dNTP, 1.25 µl of DMSO, 0.25 µl of Illumina PE primer #1, 0.25 µl of Illumina PE primer #2, 0.25 µl of Hotstart Phusion polymerase, and 2 µl of the DNA. The PCR program used was: 98°C for 2 minutes; 12 cycles of 98°C for 15 seconds, 65°C for 30 seconds, 72°C for 30 seconds; and 72°C for 5 min. DNA was purified using Agencourt AMPure XP beads (Beckman Coulter, IN) in a ratio of 1.0 to 1.0 of PCR product to beads. Exonic regions were captured in solution using the Agilent SureSelect v.4 kit (Agilent, Santa Clara, CA). The captured library was then purified with a Qiagen MinElute column purification kit and eluted in 17 µl of 70°C EB to obtain 15 µl of captured DNA library. To amplify the captured DNA library, eight 30 µL PCR reactions containing 19 µl of H_2_O, 6 µl of 5 x Phusion HF buffer, 0.6 µl of 10 mM dNTP, 1.5 µl of DMSO, 0.30 µl of Illumina PE primer #1, 0.30µl of Illumina PE primer #2, 0.30 µl of Hotstart Phusion polymerase, and 2 µl of captured exome library were set up. The PCR program used was: 98°C for 30 seconds; 14 cycles of 98°C for 10 seconds, 65°C for 30 seconds, 72°C for 30 seconds; and 72°C for 5 min. To purify PCR products, a NucleoSpin Extract II purification kit (Macherey-Nagel, PA) was used. Paired-end sequencing resulting in 100 bases from each end of the fragments was performed using Illumina HiSeq 2500 (Illumina, San Diego, CA).

Plasma Preparation: Peripheral blood was collected in K2-EDTA tubes after informed consent was obtained, and plasma was isolated as previously described^29^. cfDNA from each of the plasma samples was purified using a BioChain cfDNA Extraction Kit (BioChain, cat #K5011610) using the manufacturer’s recommended protocol.

### Processing of Next Generation Sequencing Data

Somatic mutations were identified using VariantDx custom software for identifying mutations in matched tumor and normal samples from whole exome sequencing (WES). Prior to mutation calling, primary processing of sequence data for both tumor and normal samples were performed using Illumina CASAVA software (v1.8), including masking of adapter sequences. Sequence reads were aligned against the human reference genome (version hg19) using ELAND software. Candidate somatic mutations, consisting of point mutations, insertions, and deletions were then identified using VariantDx. In brief, an alignment filter was applied to exclude quality failed reads, unpaired reads, and poorly mapped reads in the tumor. A base quality filter was applied to limit inclusion of bases with reported phred quality score > 30 for the tumor and > 20 for the normal. A mutation in the tumor was identified as a candidate somatic mutation only when (i) distinct paired reads contained the mutation in the tumor; (ii) the number of distinct paired reads containing a particular mutation in the tumor was at least 10% of the total distinct read pairs; (iii) the mismatched base was not present in >1% of the reads in the matched normal sample as well as not present in a custom database of common germline variants derived from dbSNP; (iv) the position was covered in both the tumor and normal. Mutations arising from misplaced genome alignments, including paralogous sequences, were identified and excluded by searching the reference genome.

Candidate somatic mutations were further filtered based on gene annotation to identify those occurring in protein coding regions. Functional consequences were predicted using snpEff and a custom database of CCDS, RefSeq and Ensembl annotations using the latest transcript versions available on hg19 from UCSC (https://genome.ucsc.edu/). Predictions were ordered to prefer transcripts with canonical start and stop codons and CCDS or Refseq transcripts over Ensembl when available. Finally, mutations were filtered to exclude intronic and silent changes, while retaining mutations resulting in missense mutations, nonsense mutations, frameshifts, or splice site alterations. A manual visual inspection step was used to further remove artifactual changes.

### ddPCR

Cell-free DNA was extracted using the QIAGEN circulating nucleic acid kit (Catalog# 55114). Extracted cell-free DNA was analyzed with custom designed droplet digital PrimePCR™ assays using the BioRad QX200 droplet digital PCR system to determine the number of wild-type and mutant genomic equivalents following the manufacturer’s recommendations. A mutation was selected for each tumor based on the results of the whole exome sequencing (WES) results. ddPCR was then performed on DNA derived from the plasma and ctDNA levels were quantified. These data were used to calculate the overall mutant allele frequency (MAF) for each somatic mutation, defined as the number of mutant counts divided by the total number of counts for a given amplicon.

### RealSeqS

RealSeqS uses a single primer pair to amplify about 750,000 loci scattered throughout the genome^23^. After massively parallel sequencing, gains or losses of each of the 39 chromosome arms covered by the assay were determined using a bespoke statistical learning method^30^. A support vector machine (SVM) was used to discriminate between aneuploid and euploid samples. The SVM was trained using 2651 aneuploid samples and 1348 euploid plasma samples. Samples were scored as positive when the genome wide aneuploidy score was > 0.28.

Plasma samples were also analyzed for genomic DNA contamination using RealSeqS. RealSeqS enables the detection of genomic DNA by virtue of the differently-sized amplicons generated during PCR amplification. Reads at the 1241 amplicons described in Douville *et al*. indicates the presence of genomic DNA^23^.

### Statistical Analysis Methods

Comparison of GAS at 97% specificity was conducted with a one-way ANOVA with post-hoc Tukey’s correction. Clinicopathological data were compared using a χ^2^ test or linear regression with Spearman’s correlation. A *p* ≤ 0.05 was considered significant.

## Data Availability

All data are available upon request.

## FIGURES

**Supplemental Figure 1.**
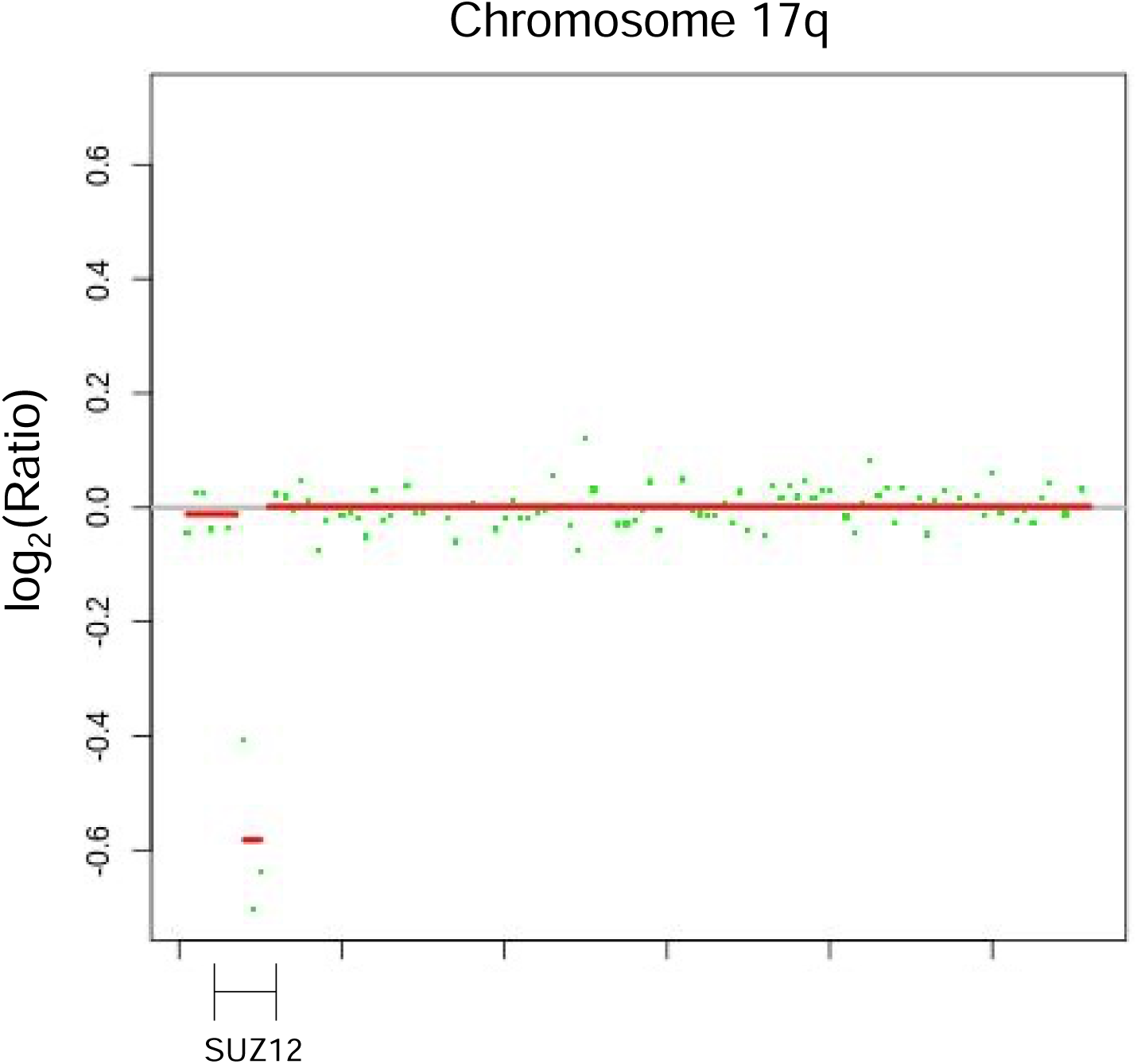
Patients with MPNST demonstrated loss of *NF1* and *SUZ12* on chromosome 17q. Each dot represents a sub-chromosomal locus covered by RealSeqS.

## SUPPLEMENTAL TABLE LEGENDS

**Supplemental Table 1**. Genome wide aneuploidy (GAS) scores and number of useable UIDs for 883 healthy controls.

**Supplemental Table 2**. Clinicopathological characteristics of the patient cohort and the results of genome wide aneuploidy analysis and mutation analysis of ctDNA

**Supplemental Table 3**. Detailed genome wide aneuploidy scores per chromosome arm

**Supplemental Table 4**. Focal copy number change analysis for 13 commonly altered loci in MPNST

**Supplemental Table 5**. Whole exome sequencing results for 6 patients with ctDNA mutation analysis

## REFERENCES

1 DeClue, J. E. et al. Abnormal regulation of mammalian p21ras contributes to malignant tumor growth in von Recklinghausen (type 1) neurofibromatosis. Cell 69, 265–273, doi:10.1016/0092-8674(92)90407-4 (1992).

2 Carroll, S. L. Molecular mechanisms promoting the pathogenesis of Schwann cell neoplasms. Acta Neuropathol 123, 321–348, doi:10.1007/s00401-011-0928-6 (2012).

3 Rasmussen, S. A., Yang, Q. & Friedman, J. M. Mortality in neurofibromatosis 1: an analysis using U.S. death certificates. Am J Hum Genet 68, 1110–1118, doi:10.1086/320121 (2001).

4 De Raedt, T. et al. Elevated risk for MPNST in NF1 microdeletion patients. Am J Hum Genet 72, 1288–1292, doi:10.1086/374821 (2003).

5 Zheng, H. et al. Induction of abnormal proliferation by nonmyelinating schwann cells triggers neurofibroma formation. Cancer Cell 13, 117–128, doi:10.1016/j.ccr.2008.01.002 (2008).

6 Yang, F. C. et al. Nf1-dependent tumors require a microenvironment containing Nf1+/-- and c-kit-dependent bone marrow. Cell 135, 437–448, doi:10.1016/j.cell.2008.08.041 (2008).

7 Zhu, Y., Ghosh, P., Charnay, P., Burns, D. K. & Parada, L. F. Neurofibromas in NF1: Schwann cell origin and role of tumor environment. Science 296, 920–922, doi:10.1126/science.1068452 (2002).

8 Cichowski, K. et al. Mouse models of tumor development in neurofibromatosis type 1. Science 286, 2172–2176, doi:10.1126/science.286.5447.2172 (1999).

9 De Raedt, T. et al. PRC2 loss amplifies Ras-driven transcription and confers sensitivity to BRD4-based therapies. Nature 514, 247–251, doi:10.1038/nature13561 (2014).

10 Legius, E. et al. TP53 mutations are frequent in malignant NF1 tumors. Genes Chromosomes Cancer 10, 250–255, doi:10.1002/gcc.2870100405 (1994).

11 Perry, A. et al. Differential NF1, p16, and EGFR patterns by interphase cytogenetics (FISH) in malignant peripheral nerve sheath tumor (MPNST) and morphologically similar spindle cell neoplasms. J Neuropathol Exp Neurol 61, 702–709, doi:10.1093/jnen/61.8.702 (2002).

12 Zhang, M. et al. Somatic mutations of SUZ12 in malignant peripheral nerve sheath tumors. Nat Genet 46, 1170–1172, doi:10.1038/ng.3116 (2014).

13 Mautner, V. F. et al. Assessment of benign tumor burden by whole-body MRI in patients with neurofibromatosis 1. Neuro Oncol 10, 593–598, doi:10.1215/15228517-2008-011 (2008).

14 Higham, C. et al. Atypical neurofibromas in neurofibromatosis 1 (NF1): Clinical, imaging and pathologic characteristics. Journal of Clinical Oncology 34, 11035–11035, doi:10.1200/JCO.2016.34.15_suppl.11035 (2016).

15 Derlin, T. et al. Comparative effectiveness of 18F-FDG PET/CT versus whole-body MRI for detection of malignant peripheral nerve sheath tumors in neurofibromatosis type 1. Clin Nucl Med 38, e19–25, doi:10.1097/RLU.0b013e318266ce84 (2013).

16 Ferner, R. E. et al. [18F]2-fluoro-2-deoxy-D-glucose positron emission tomography (FDG PET) as a diagnostic tool for neurofibromatosis 1 (NF1) associated malignant peripheral nerve sheath tumours (MPNSTs): a long-term clinical study. Ann Oncol 19, 390–394, doi:10.1093/annonc/mdm450 (2008).

17 Azizi, A. A. et al. Monitoring of plexiform neurofibroma in children and adolescents with neurofibromatosis type 1 by [(18) F]FDG-PET imaging. Is it of value in asymptomatic patients? Pediatr Blood Cancer 65, doi:10.1002/pbc.26733 (2018).

18 Reinert, C. P. et al. Comprehensive anatomical and functional imaging in patients with type I neurofibromatosis using simultaneous FDG-PET/MRI. Eur J Nucl Med Mol Imaging 46, 776–787, doi:10.1007/s00259-018-4227-5 (2019).

19 Pekmezci, M. et al. Morphologic and immunohistochemical features of malignant peripheral nerve sheath tumors and cellular schwannomas. Mod Pathol 28, 187–200, doi:10.1038/modpathol.2014.109 (2015).

20 Fletcher, C. D. The evolving classification of soft tissue tumours - an update based on the new 2013 WHO classification. Histopathology 64, 2–11, doi:10.1111/his.12267 (2014).

21 Hirbe, A. C. et al. beta-III-spectrin immunohistochemistry as a potential diagnostic tool with high sensitivity for malignant peripheral nerve sheath tumors. Neuro Oncol 20, 858–860, doi:10.1093/neuonc/noy038 (2018).

22 Brohl, A. S., Kahen, E., Yoder, S. J., Teer, J. K. & Reed, D. R. The genomic landscape of malignant peripheral nerve sheath tumors: diverse drivers of Ras pathway activation. Sci Rep 7, 14992, doi:10.1038/s41598-017-15183-1 (2017).

23 Douville, C. et al. Assessing aneuploidy with repetitive element sequencing. Proc Natl Acad Sci U S A 117, 4858–4863, doi:10.1073/pnas.1910041117 (2020).

24 Ferner, R. E. & Gutmann, D. H. International consensus statement on malignant peripheral nerve sheath tumors in neurofibromatosis. Cancer Res 62, 1573–1577 (2002).

25 Akshintala, S. et al. Longitudinal evaluation of peripheral nerve sheath tumors in neurofibromatosis type 1: growth analysis of plexiform neurofibromas and distinct nodular lesions. Neuro Oncol 22, 1368–1378, doi:10.1093/neuonc/noaa053 (2020).

26 Canavese, F. & Krajbich, J. I. Resection of plexiform neurofibromas in children with neurofibromatosis type 1. J Pediatr Orthop 31, 303–311, doi:10.1097/BPO.0b013e31820cad77 (2011).

27 Reilly, K. M. et al. Neurofibromatosis Type 1-Associated MPNST State of the Science: Outlining a Research Agenda for the Future. J Natl Cancer Inst 109, doi:10.1093/jnci/djx124 (2017).

28 Bettegowda, C. et al. Exomic sequencing of four rare central nervous system tumor types. Oncotarget 4, 572–583, doi:10.18632/oncotarget.964 (2013).

29 Diehl, F. et al. Circulating mutant DNA to assess tumor dynamics. Nat Med 14, 985–990, doi:10.1038/nm.1789 (2008).

30 Douville, C. et al. Detection of aneuploidy in patients with cancer through amplification of long interspersed nucleotide elements (LINEs). Proc Natl Acad Sci U S A 115, 1871–1876, doi:10.1073/pnas.1717846115 (2018).

